# Frequency of routine testing for COVID-19 in high-risk healthcare environments to reduce outbreaks

**DOI:** 10.1101/2020.04.30.20087015

**Authors:** Elizabeth T Chin, Benjamin Q Huynh, Lloyd A. C. Chapman, Matthew Murrill, Sanjay Basu, Nathan C Lo

**Author notes:** Co-first author (Elizabeth T Chin, Benjamin Q Huynh). **Correspondence:**, Nathan C. Lo, MD PhD, Department of Medicine University of California, San Francisco, San Francisco, CA 94110 USA.

## Abstract

Abstract

Routine asymptomatic testing strategies for COVID-19 have been proposed to prevent outbreaks in high-risk healthcare environments. We used simulation modeling to evaluate the optimal frequency of viral testing. We found that routine testing substantially reduces risk of outbreaks, but may need to be as frequent as twice weekly.

## Introduction

Outbreaks of SARS-CoV-2, the causative virus of COVID-19, have been commonly documented in high-risk healthcare environments ranging from skilled nursing facilities and hospitals to homeless shelters.^1,2^ Routine viral testing strategies with polymerase chain reaction (PCR) of asymptomatic persons have been proposed to detect and prevent outbreaks in high-risk healthcare environments, by testing residents and workers at routine intervals in absence of documented cases. Yet it remains unclear how often routine asymptomatic testing would need to be performed, and how effective such a strategy would be to prevent outbreaks of COVID-19. The United States Centers for Disease Control and Prevention has recently issued partial guidance for viral testing during an outbreak, although no preventive testing guidelines exist.^3,4^ In this study, we aimed to estimate the effectiveness of routine testing with PCR to reduce transmission of COVID-19.

## Methods

### Overview

We developed a simulation model of SARS-CoV-2 transmission to evaluate the effectiveness of various frequencies of routine PCR testing of all persons in a high-risk healthcare environment (i.e., long term residents or patients admitted to hospitals, daily healthcare workers) to reduce cases of COVID-19. Some examples of representative healthcare environments include nursing facilities, hospitals, clinics, dialysis centers, and substance use treatment centers. The primary study outcome for each strategy was the simulated reduction in the mean control reproduction number (R_c_), corresponding to the average number of secondary infections caused by an infected person averaged over the simulation period, starting with a fully susceptible population, and accounting for the impact of interventions. For interpretation, a mean control reproduction number below one would ensure decline in the number of cases when averaged over time.

### Model structure

The SARS-CoV-2 transmission model was a stochastic microsimulation, where individuals were simulated and assigned a health state that included being susceptible to infection (non-immune), early infectious, late infectious, or recovered and immune (Figure A1). We simulated transmission in a population of 100 people within a healthcare environment interacting with a community with daily incidence of 0.5%, over 10 months, where people in the healthcare environment were constantly interacting with new community members. We chose a high daily incidence to ensure sufficient number of new infections for the simulation; this choice should not affect the study results, and was also tested in sensitivity analysis. We used published data on the natural history of SARS-CoV-2, including an estimated 5-day incubation period and 9-day infectious period. We inferred the probability of infection per day of work based on the estimated infectiousness profile of SARS-CoV-2 (including infectiousness beginning 4 days prior to onset of symptoms) (Figure A2).^5^ We assumed a 40% sub-clinical proportion, with 50% relative infectiousness of sub-clinical infections to clinically apparent cases.^1,6,7^ More details on the model structure and parameters are available in the Appendix.

### Simulation

We modeled transmission occurring within a high-risk healthcare environment that was fully susceptible through introduction from the community. We assumed a basic reproduction number (R_0_) within the healthcare environment corresponding to the number of secondary infections caused by an infected person in an entirely susceptible population in absence of intervention.^5,8^ We tested a base case R_0_ of 2.5 based on published literature, but also varied R_0_ to test lower values that may represent complementary interventions (e.g. universal masking, social distancing).

We evaluated routine asymptomatic PCR testing of various frequencies, from daily to once monthly testing. We modeled the sensitivity of PCR testing as a function of day of infection based on data of time-varying sensitivity of this test modality (50-80% during first two weeks; Figure A3)^9^, and PCR specificity as 98-100%. We estimated the effect of testing on R_c_, with a goal of achieving a R_c_ below one. We assumed that persons self-isolated upon symptom onset, and persons with PCR-confirmed infection self-isolated one day after being tested, while those that were not detected remained in the environment and potentially infected others. We performed Monte Carlo sampling across the uncertainty ranges of each parameter to estimate the range of possible outcomes. We performed sensitivity analysis by varying test result delays and test performance. Additional details are available in the Appendix and data and code are available online (https://github.com/etchin/covid-testing).

## Results

In this microsimulation, with daily testing in high-risk environments by PCR and an assumed basic reproduction number R_0_ of 2.5, we estimated an 82.2% (95% CI: 82.0-82.5) reduction in R_c_, corresponding to R_c_=0.44. When testing persons every three days, we observed a 61.4% (95% CI: 61.2-61.7) reduction, corresponding to R_c_=0.97. When testing weekly, we observed a 36.9% (95% CI: 36.5-37.2) reduction, corresponding to R_c_=1.58; and when testing monthly, we observed a 8.9% (95% CI: 8.7-9.2) reduction, corresponding to R_c_=2.28 (Table A3).

The optimal testing frequency to bring R_c_ below one was dependent on baseline R_0_ (Figure 1). In an environment with R_0_ = 2.5,^5,8^ testing would have to occur almost every other day to bring R_c_ below one. If R_0_ = 2,^5,8^ testing would need to occur at least twice weekly (every 3-4 days), unless other measures were added to testing and self-isolation. If assuming R_0_ = 1.5, testing weekly would suffice.

**Figure 1:**
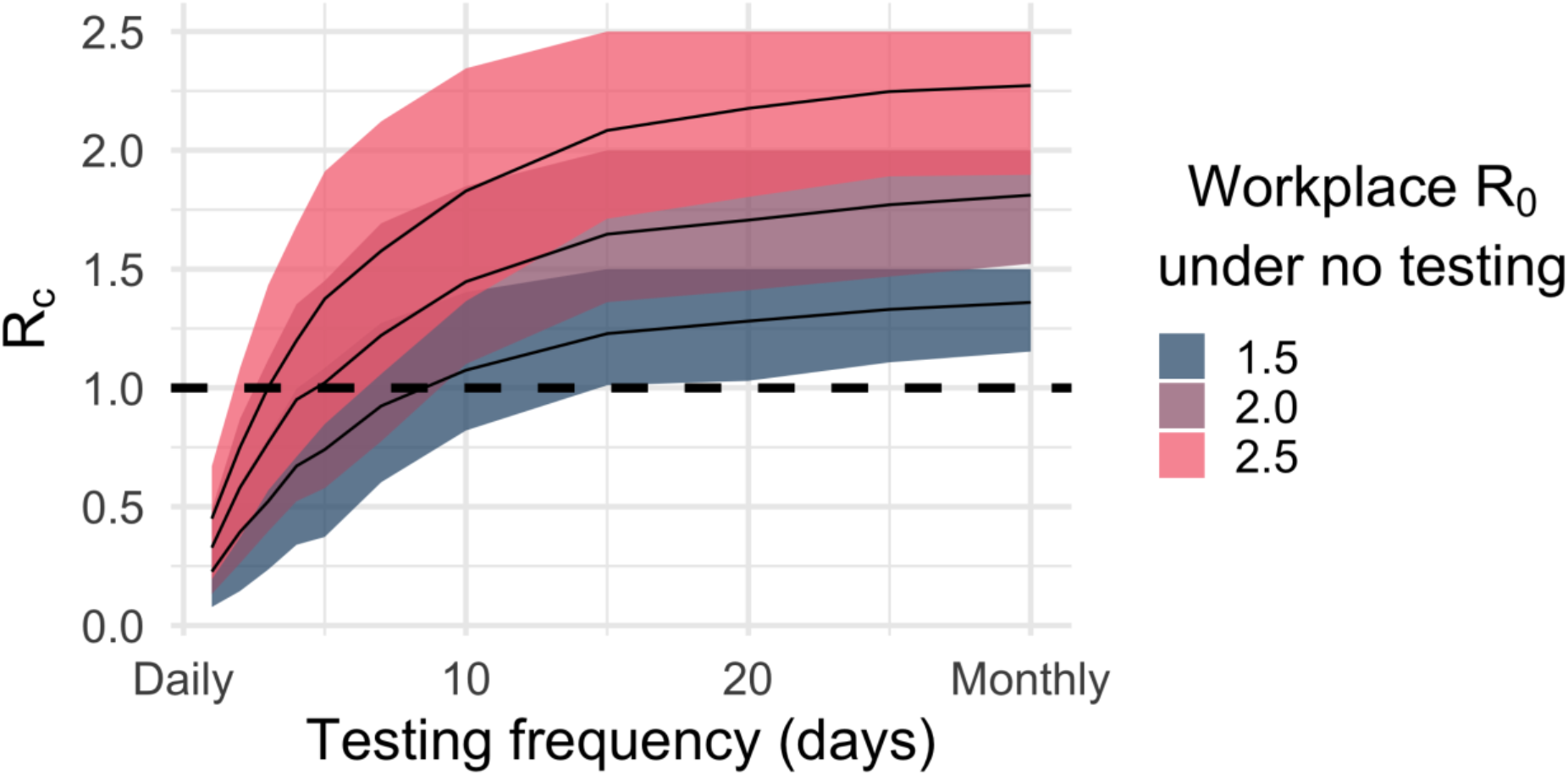
Projected impact of routine PCR testing frequency on the mean control reproduction number under different testing scenarios. We estimated the effectiveness of increasing frequency of routine PCR testing to reduce the mean control reproduction number, R_c_, under different assumptions on the underlying basic reproduction number, R_0_. The x-axis refers to the frequency of PCR testing simulated, from daily (testing frequency of 1 day) to once a month (testing frequency of 30 days). The y-axis represents the mean control reproduction number (R_c_), which is the average number of secondary infections caused by an infected person averaged over the simulation period, starting with a fully susceptible population, and accounting for the impact of interventions. The goal is to reduce R_c_ to below one to ensure decline in the number of cases when averaged over time. Bands represent the interquartile range accounting for parameter and stochastic uncertainty.

In sensitivity analysis, we observed only small changes in results with variation in test sensitivity, but large changes with variation in test result delays. With R_0_=1.5, reducing test sensitivity by 20% reduced the impact of daily testing (in terms of reduction in R_c_) from 85.3% (95% CI: 85.1-85.6) to 80.7% (95% CI: 8o.5-81.0). Longer test result delays of 3 and 5 days reduced daily testing impact from 85.3% reduction in R_c_ to 56.5% (95% CI: 56.2-56.9) and 25.9% (95% CI: 25.4-26.3), respectively. In an ideal case with zero delay and perfect sensitivity, daily testing reduced R_c_ by 98.9% (95% CI: 98.6-99.1) (Figure A4). Varying the backgrournd incidence had minimal impact on the study results (Figure A5).

## Discussion

This simulation study finds that in high-risk settings with ongoing community-based transmission, frequent (twice-weekly) routine asymptomatic viral testing may be required to prevent outbreaks and reduce case counts of COVID-19. Due to the imperfect sensitivity of PCR testing and infectiousness early in the natural history, even with frequent testing, a meaningful proportion of infected persons may be missed. We find that strategies with less frequent testing - such as once-a-week testing - may be sufficient in settings with low community incidence, especially when implemented with additional infection control measures. Furthermore, we find that delays in returning test results would severely impact the effectiveness of routine testing strategies.

The study conclusions are most applicable to high-risk healthcare environments, with long-term residents and daily workers. These settings include nursing facilities, hospitals, prisons, homeless shelters, dialysis centers, and other healthcare and non-healthcare environments. The assumptions in the model are most applicable in a setting with ongoing community transmission of SARS-CoV-2, as evidenced by ongoing new infections. In settings with higher community incidence, testing multiple times per week would be required to prevent an outbreak and control case counts, and require the addition of other control strategies (e.g. universal masking, social distancing). Our study conclusions are similar to recently published model-based analyses on PCR testing strategies,^10,11^ which support the finding that very frequent testing (every 2-3 days) is required to have a meaningful impact on transmission, despite modeling different environments.

The study has limitations in the model assumptions and available data. Transmission of SARS-CoV-2 is documented to have high degree of heterogeneity across settings, whereas we used a transmission rate that considered an average among high-incidence settings such as nursing facilities. Our analysis focused on outbreaks and transmission in high-risk environments, rather than the population at large. Furthermore, routine PCR testing would require substantial resources, logistical support, and high participation from the population, with consideration of cost-effectiveness.^12^ We assumed that results of testing would be available after one day which may only be possible in higher resource settings, but also tested the impact of slower turnaround time, which reduced the overall effectiveness of this strategy.

In conclusion, our findings support that routine testing strategies can provide benefit to reduce transmission in high-risk environments with frequent testing but may require complementary strategies to reliably prevent outbreaks of COVID-19. Further evidence should be generated on the use of strategies in combination with testing, including masking, ventilation changes, disinfection, and physical distancing.^8,13^

## Contributions

E.T.C, B.Q.H, L.A.C.C. and N.C.L developed the transmission model. E.T.C, N.C.L, and B.Q.H coded the simulation and analysis. S.B. and N.C.L. supervised the study. All authors contributed to study design, interpretation of results, and writing of the manuscript.

## Data Availability

Data and code available at: https://github.com/etchin/covid-testing.

## Acknowledgments

E.T.C acknowledges support by the National Science Foundation Graduate Research Fellowship under Grant No. DGE 1656518. NCL and LACC are supported by the University of California, San Francisco. B.Q.H acknowledges support by the National Science Foundation Graduate Research Fellowship under Grant No. DGE 1656518 and the National Library of Medicine under Training Grant T15 LM 007033. Funding sources had no role in the writing of this correspondence or the decision to submit for publication.

## Declaration of interests

SB serves on the City of San Francisco’s Department of Public Health street outreach team for homeless adults affected by COVID-19, as a provider at the HealthRight360 Integrated Care Center, and as an employee of Collective Health, all of which provide COVID-19 testing. The views expressed here reflect the opinions of the authors and not necessarily those of affiliated organizations.

## Technical Appendix

### Methods

We developed a microsimulation model of SARS-CoV-2 transmission to estimate the impact of variable frequency of routine asymptomatic PCR testing on the mean control reproduction number, R_c_, in a high-risk healthcare environment. We developed a susceptible-exposed-infectious-recovered (SEIR)-like model in which individuals interact with an age-structured community population as well as within a high-risk healthcare environment (e.g. nursing home, hospital). We simulated PCR-based testing for each individual in a healthcare environment and varied the time intervals for routine PCR testing from daily to monthly. We assumed persons self-isolate when receiving a positive test or when symptoms occur, so that transmission within the healthcare environment occurs only from sub-clinical or early-clinical infected persons. We also assumed individuals take one day to receive results after testing, although we varied this is in a sensitivity analysis. We probabilistically varied the following parameters: incubation time, early infectious period, late infectious period, test sensitivity, and test specificity (see Table A2).

The model tracked three features of each simulated person: (i) the person’s true state of infection (susceptible, early sub-clinical infection, late sub-clinical infection, early clinical infection, late clinical infection, or recovered) (Figure A1, Table A1); (ii) the observed state of infection based on test results (uninfected, currently infected based on positive PCR, or immune based on positive PCR followed by completion of a 14 day self-isolation period); and (iii) whether the person was present in the healthcare environment.

A susceptible individual can be exposed to infectious individuals in both the community and other members of the healthcare environment. We applied a constant probability of infection from the community, where incidence was assumed to be 0.5%. We chose a high daily incidence to ensure sufficient number of new infections for the simulation; this choice should not affect the study results (mean control reproduction number), and was also tested in sensitivity analysis to verify this (Figure A5). Simulations using a lower incidence (0.1%, 0.01%, and 0.001%) affected the precision of the estimate, but the mean remained stable.

We assume a healthcare setting where the majority (75%) of contacts occur between a person in the healthcare environment and the community. We assume that 100% of symptomatic individuals in the healthcare environment and 75% of the community will self-isolate. The force of infection, *λ_i_(t*), on each individual *i* on day *t* is proportional to the prevalence of infectious individuals within the community and healthcare environment they are exposed to on day *t* and their infectiousness:

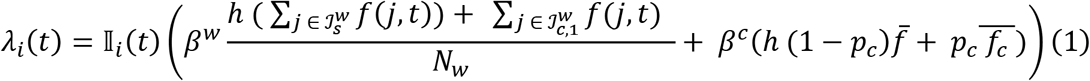

Here, *β* is the transmission rate coefficient derived from the basic reproductive number - healthcare environment and community transmission rate coefficients being denoted by superscripts w and c respectively. 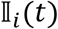 is an indicator function for whether the individual is present in the healthcare environment on day *t*. 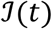 represents the infectious individuals on day *t* in different states of infection - subclinical and clinical, denoted by subscripts s and *c* respectively, early and late stage, denoted by subscripts 1 and 2 respectively. *h* is the infectiousness of subclinically infected individuals relative to those with clinical symptoms. *f* is a function for the daily infectiousness of an individual *j* on day *t* (Figure A2). 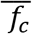is the mean daily infectiousness across all days of infection and 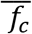 is the mean daily infectiousness for clinical infections in the community, where we assume that 75% of symptomatic individuals self-isolate. See Table A2 for parameter values used in the simulations.

Each individual believed to be uninfected in the population is tested at varying intervals. We simulated transmission among 100 healthcare workers for 300 days across 1000 simulations for each parameter setting. We model three risk groups - low: R_0_ = 1.5, medium: R_0_ = 2, and high: R_0_ = 2.5 - under time-varying PCR sensitivities (Figure A3) with a test result delay of one day. To model the ideal case, we assume 100% sensitivity and no delay in test results in a low-risk group. To calculate the reduction in transmission to estimate R_c_, we take the mean total number of infectious days, weighted by infectiousness (Table A3), under a specific testing frequency and divide it by the counterfactual, which we define as the mean total number of weighted infectious days under no testing.

To estimate R_c_, we multiplied the R_0_ for the workplace under no testing by the reduction in transmission at a given testing frequency. The bands in Figure 1 represent the interquartile range of the distribution of the effective reproduction number over uncertainty in the parameter values and stochasticity. The model assumes a constant population and that individuals gain immunity in the short-term after recovery.

We additionally modeled lower sensitivity rapid tests, such as antigen tests, by estimating the reduction in infectiousness and cumulative number of infections under lower test sensitivity, using a discount factor of 0.8, and zero and one-day delays in test results. For further sensitivity analysis, we also modeled test result delays of 3 and 5 days to assess the effect of test turnaround times on testing impact (Figure A4).

Data and code available at: https://github.com/etchin/covid-testing.

**Figure A1.**
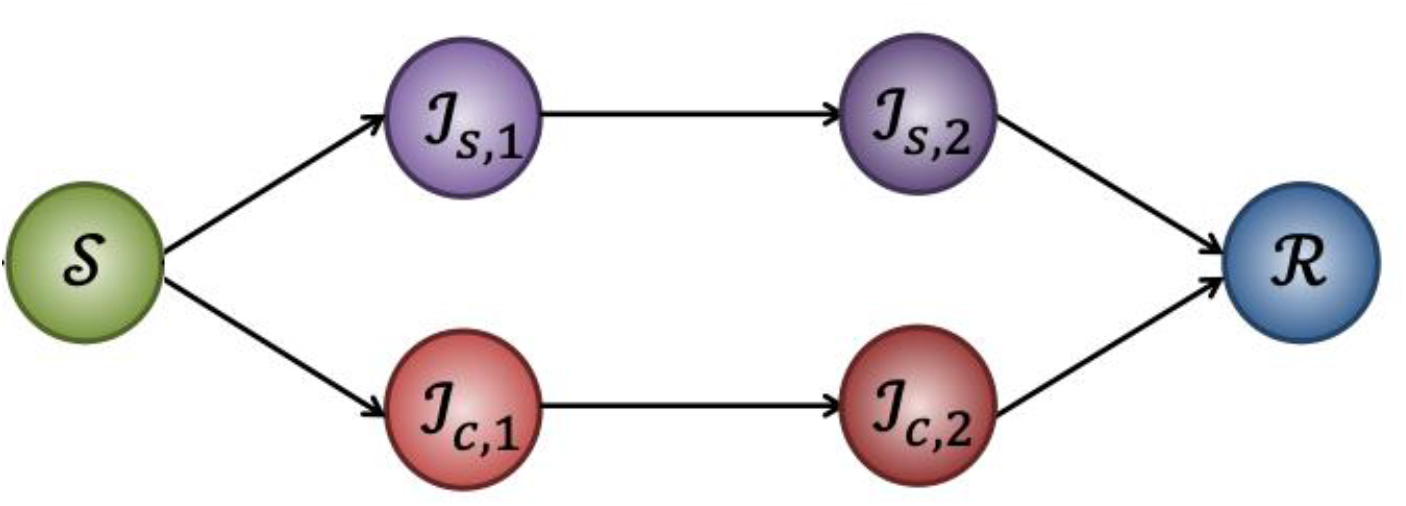
Structure of stochastic individual-level model of COVID-19 transmission. The labels of each state correspond with definitions in Table A1.

**Table A1:**
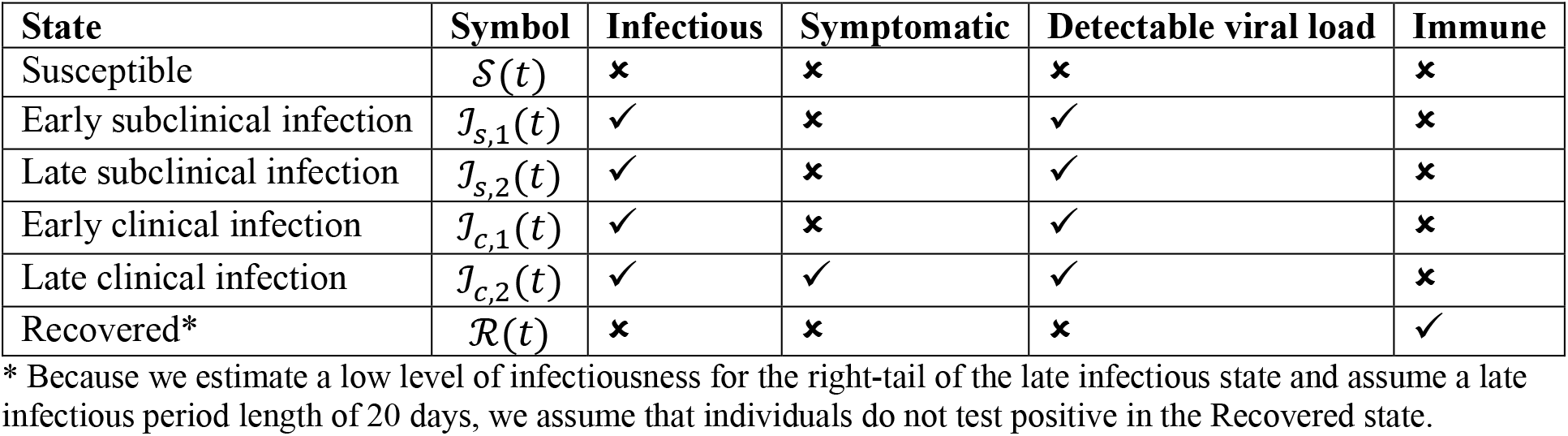
Definition of states in the transmission model.

| State | Symbol | Infectious | Symptomatic | Detectable viral load | Immune |
| --- | --- | --- | --- | --- | --- |
| Susceptible | $\mathcal{S}(t)$ | ✗ | ✗ | ✗ | ✗ |
| Early subclinical infection | $\mathcal{I}_{s,1}(t)$ | ✓ | ✗ | ✓ | ✗ |
| Late subclinical infection | $\mathcal{I}_{s,2}(t)$ | ✓ | ✗ | ✓ | ✗ |
| Early clinical infection | $\mathcal{I}_{c,1}(t)$ | ✓ | ✗ | ✓ | ✗ |
| Late clinical infection | $\mathcal{I}_{c,2}(t)$ | ✓ | ✓ | ✓ | ✗ |
| Recovered* | $\mathcal{R}(t)$ | ✗ | ✗ | ✗ | ✓ |

\* Because we estimate a low level of infectiousness for the right-tail of the late infectious state and assume a late infectious period length of 20 days, we assume that individuals do not test positive in the Recovered state.

**Table A2:** Model parameters and distributions in model.

| Parameter | Distribution/Value | References |
| --- | --- | --- |
| Sensitivity of PCR | Time-varying* estimates fit to a truncated normal distribution by day of infection (Figure A3) | 1 |
| Specificity of PCR | 98-100% fit to a truncated normal distribution | 2-5 |
| Infectiousness | Time-varying estimates by day of infection. Fit to a gamma distribution Infectiousness is assumed to peak at the start of the late infectious period fit to a truncated normal distribution (Figure A2) | 6 |
| Early infectious period | 5.2 days (95% CI, 4.1-6.4) fit to a log-normal distribution | 6 |
| Late infectious period | Up to 20 days, infectiousness decreases after symptom onset (Figure A2) | 6 |
| Proportion sub-clinical | 40% | 7 |
| Discount factor for sub-clinical infectiousness | 50% as infectious as symptomatic infection | 8,9 |

\* Sensitivity estimates by day of infection were obtained from Kucirka et al. [1]. Sensitivity estimates during the incubation period were excluded because sensitivity was fit using the data of only one individual from the Danis et al study (PMID: 32277759). This patient tested negative for SARS-CoV-2 when obtained using nasopharyngeal swabs during the pre-symptomatic phase. Upon symptom onset, the patient had an endotracheal aspirate (ETA) sample test positive; however, the nasopharyngeal swabs of the same day and the following days remained negative. Thus, we assume exponential growth of test sensitivity during the early infectious period.

**Table A3:** Ranges of mean percent reduction of infectiousness in a healthcare environment.

| Testing frequency | Low Risk ($R_0 = 1.5$) | Medium Risk ($R_0 = 2$) | High Risk ($R_0 = 2.5$) | Ideal Case |
| --- | --- | --- | --- | --- |
| 1 | 85.3% (85.1,85.6) | 83% (82.8,83.2) | 82.2% (82,82.5) | 98.9% (98.6,99.1) |
| 2 | 73.9% (73.6,74.2) | 71.5% (71.3,71.8) | 69.5% (69.2,69.7) | 94.8% (94.6,95.1) |
| 3 | 64.5% (64.2,64.8) | 62.3% (62,62.6) | 61.4% (61.2,61.7) | 89.6% (89.4,89.9) |
| 4 | 56.6% (56.2,56.9) | 53.2% (53,53.5) | 52.1% (51.8,52.4) | 85.1% (84.8,85.3) |
| 5 | 49.7% (49.4,50.1) | 47.4% (47.1,47.7) | 45.4% (45.1,45.7) | 78.3% (78.1,78.6) |
| 7 | 38.1% (37.8,38.5) | 35.9% (35.6,36.3) | 36.9% (36.5,37.2) | 69.5% (69.2,69.8) |
| 10 | 30.5% (30.2,30.9) | 27.8% (27.5,28.1) | 27.4% (27.1,27.7) | 61.5% (61.2,61.8) |
| 15 | 15.6% (15.2,15.9) | 17.9% (17.6,18.2) | 16.8% (16.5,17.1) | 40.6% (40.3,41) |
| 20 | 12% (11.7,12.4) | 12.7% (12.4,12.9) | 12.6% (12.3,12.9) | 44.2% (43.9,44.5) |
| 25 | 9.4% (9,9.7) | 9.3% (9.1,9.6) | 12.3% (12,12.6) | 40.4% (40,40.7) |
| 30 | 8.8% (8.5,9.1) | 8.2% (7.9,8.5) | 8.9% (8.7,9.2) | 31% (30.7,31.4) |

Simulations were stratified for various risk groups (low: $R_0 = 1.5$ , medium: $R_0 = 2$ , high: $R_0 = 2.5$ ) under time-varying PCR test sensitivities with a test result delay of 1 day. The percent reduction under the ideal case was simulated with 100% sensitivity and no test result delay in a low-risk population.

**Figure A2:**
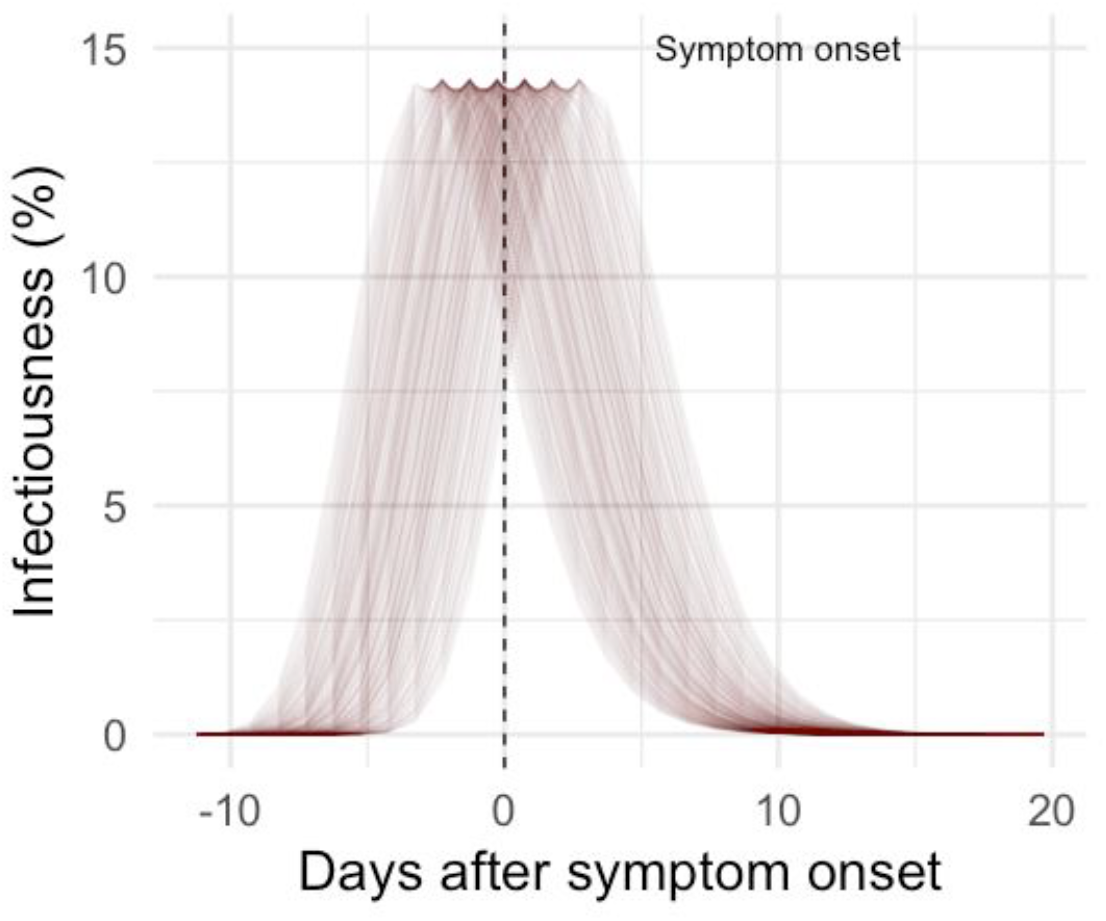
Infectiousness by day of infection.

**Figure A3.**
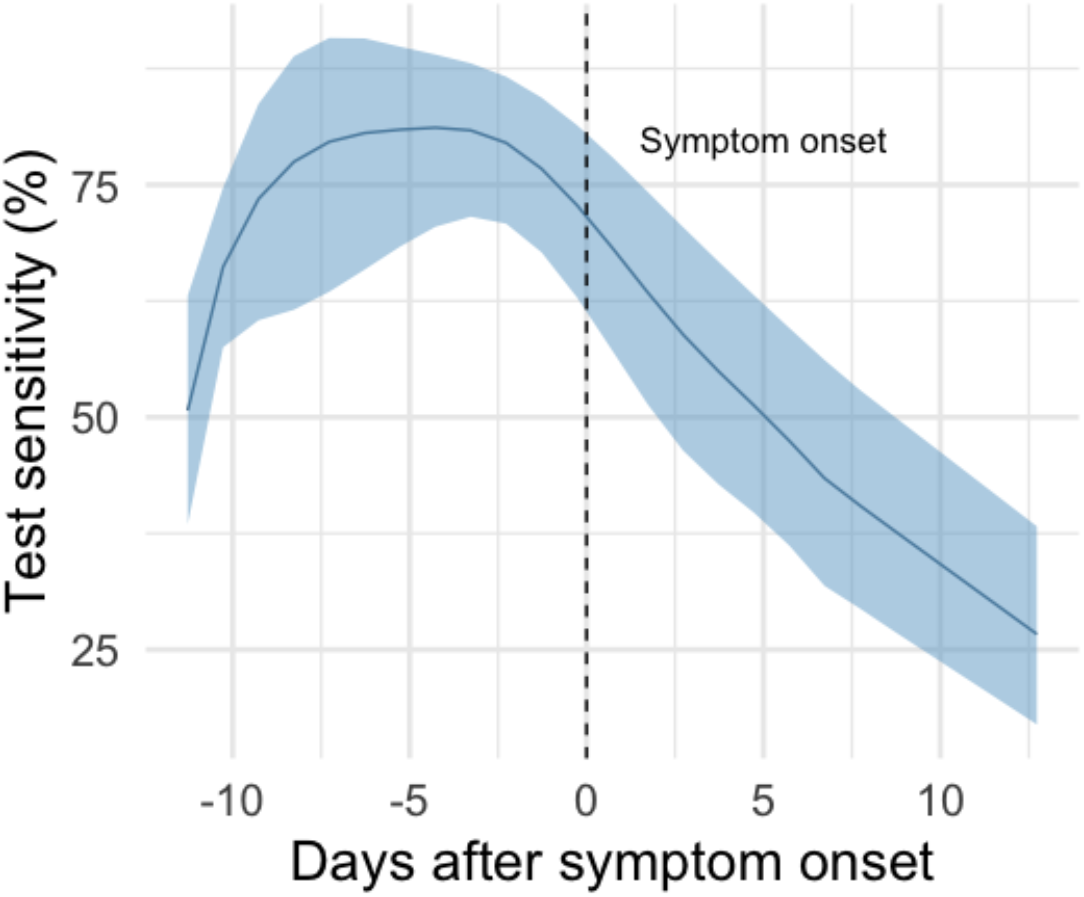
PCR sensitivity by day of infection.

**Figure A4:**
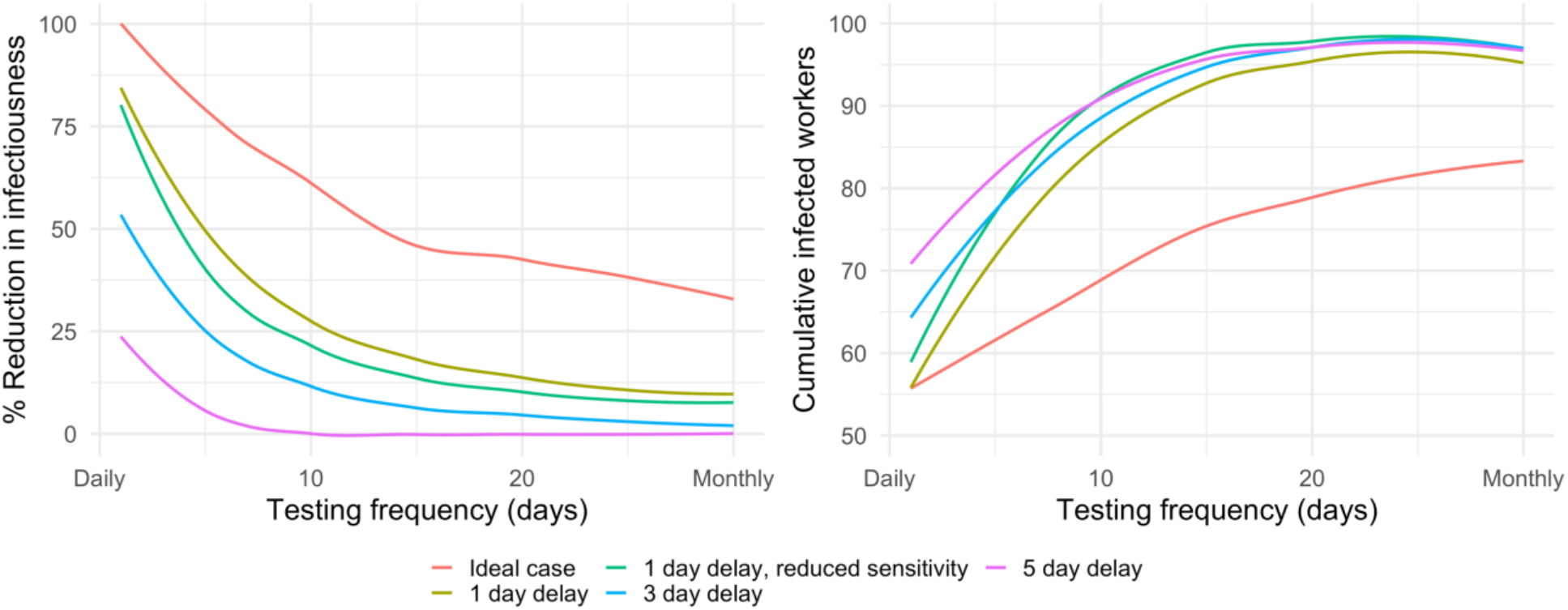
Reduction in infectiousness and cumulative infected workers under various testing strategies.

**Figure A5:**
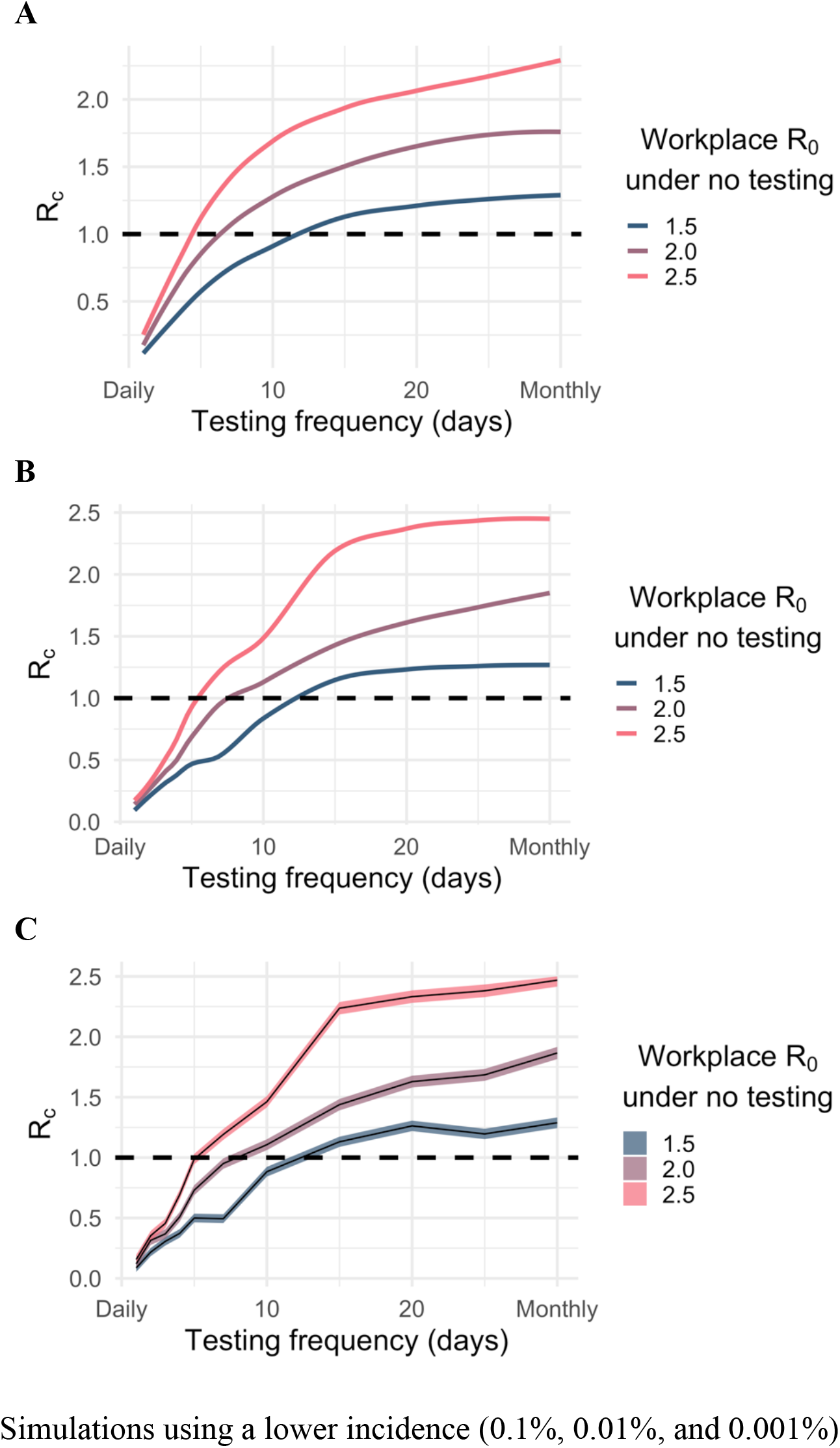
Projected impact of routine PCR testing frequency on the mean control reproduction number under different daily incidence. We repeated the base case simualation while testing alternate daily incidence estimates including: (A) 0.1%; (B) 0.01%; and (C) 0.001%.

## Notes

### Competing Interest Statement

The authors have declared no competing interest.

### Summary of Updates

Clarification on wording and additional sensitivity analyses.

